# Specific Dynamic Variations in the Peripheral Blood Lymphocyte Subsets in COVID-19 and Severe Influenza A Patients: A Retrospective Observational Study

**DOI:** 10.1101/2020.08.15.20175455

**Authors:** Fang Qian, Guiju Gao, Yangzi Song, Yanli Xu, Aibin Wang, Sa Wang, Yiwei Hao, Meiling Chen, Xiaoyang Ma, Tianwei Zhao, Xiaodi Guo, Zhihai Chen, Fujie Zhang

**Affiliations:** Clinical and Research Center of Infectious Diseases, Beijing Ditan Hospital, Capital Medical University; Department of Oncology, Beijing Ditan Hospital, Capital Medical University; Department of Neurology, Beijing Ditan Hospital, Capital Medical University; Department of Medical Records and Statistics, Beijing Ditan Hospital, Capital Medical University

**Keywords:** Coronavirus disease-19 (COVID-19), Severe influenza A, Lymphocyte, T cells subsets.

## Abstract

**Background:** Both COVID-19 and influenza A contribute to increased mortality among the elderly and those with existing comorbidities. Changes in the underlying immune mechanisms determine patient prognosis. This study aimed to analyze the role of lymphocyte subsets in the immunopathogenesisof COVID-19 and severe influenza A, and examined the clinical significance of their alterations in the prognosis and recovery duration.

**Methods:** By retrospectively reviewing of patients in four groups (healthy controls, severe influenza A, non-severe COVID-19 and severe COVID-19) who were admitted to Ditan hospital between 2018 to 2020, we performed flow cytometric analysis and compared the absolute counts of leukocytes, lymphocytes, and lymphocyte subsets of the patients at different time points (weeks 1- 4).

**Results:** We reviewed the patients’ data of 110 healthy blood donors, 80 Non-severe-COVID-19, 19 Severe-COVID-19 and 43 severe influenza A. We found total lymphocytes (0.93 ×10^9^/L, 0.84 ×10^9^/L vs 1.78 ×10^9^/L, *P* < 0.0001) and lymphocyte subsets (T cells, CD4^+^ and CD8^+^ T cell subsets) of both severe patients to be significantly lower than those of healthy donors at early infection stages. Further, significant dynamic variations were observed at different time points (weeks 1–4).

**Conclusions:** Our study indicates lymphopenia to be associated with disease severity and suggests the plausible role of lymphocyte subsets in disease progression, which in turn affects prognosis and recovery duration in patients with severe COVID-19 and influenza A.

## Background

Since the time the World Health Organization (WHO) declared the infection caused by the severe acute respiratory syndrome coronavirus 2 (SARS-CoV-2), Coronavirus Disease 2019 (COVID-19), a global pandemic, has seriously threatened to global public health and is still expanding. The clinical manifestations of COVID-19 can be mild or severe, which the incidence of mortality is higher among the elderly and among those with pre-existing comorbidities, including hypertension, cardiovascular and cerebrovascular disease, and diabetes, similar to influenza.[1] In addition, their modes of transmission are by contact, droplets, or fomites.[2,3] While relatively little is known about the immunopathological aspects of SARS-CoV-2 virus, there exists vast literature on the possible underlying mechanisms of disease pathogenesis of Influenza A virus.

As previous studies demonstrated that infection of humans with influenza A virus leads to an induction of apoptosis of a portion of CD3^+^, CD4^+^, CD8^+^, and CD19^+^ lymphocytes, thus resulting in a severe transient leukopenia, and that lymphocyte apoptosis, which represents a part of an overall beneficial immune response could be a possible mechanism of disease pathogenesis.[4,5]

Although the precise immunopathogenesis of COVID-19 is unknown, genome studies and standard blood investigations suggest involvement of the immune system. Previous studies have shown that severe leukopenia and lymphopenia occur during early stages of infection and subsequently become less prominent as the disease progresses in patients with severe COVID-19.[6,7]Furthermore, as lymphopenia developed in severe COVID-19 cases, it was shown to be a predictor of prognosis and a reliable indicator of disease severity and hospitalization in COVID-19 patients.[8] However, data on kinetic alterations in lymphocytes after COVID-19 infection are sparse. To the best of our knowledge, there are no comparative studies focusing on the dynamic variations in lymphocyte subsets of patients with influenza A and SARS-Cov-2 infections. Whether changes in the underlying immune mechanisms determine patient prognosis and recovery in COVID-19 remains unclear. Therefore, in view of the above, in this retrospective study, we analyzed the role of lymphocytes and lymphocyte subsets in the immunopathogenesis of COVID-19 and severe influenza A, to provide us with valuable insights for better understanding of the underlying immune mechanisms following SARS-Cov-2 and severe influenza A infections.

## Methods

### Study design and participants

We retrospectively reviewed the clinical data of all patients who were confirmed to have COVID-19 and were admitted to the Beijing Ditan Hospital from January 20, 2020 to March 17, 2020. In addition, all patients with severe influenza A who were admitted to Ditan hospital between 2018 to 2020 were recruited for this study. A group of healthy blood donors who were previously recruited in 2018 were used to provide data for the healthy controls in this study. This study was approved by the Ethics Committee of Beijing Ditan Hospital (No. 202000601). The need for individual consent was waived because of the retrospective nature of the study.

The patients with COVID-19 were divided into non-severe and severe pneumonia groups according to the Diagnosis and Treatment Protocols for Patients with COVID-19 (Trial Version 7, Revised).[9] The diagnostic criteria for patients with non-severe COVID-19 included: (1) Epidemiological history, (2) Fever or other respiratory symptoms, (3) Typical computed tomography (CT) image abnormities of viral pneumonia, and (4) Positive result of reverse transcription-polymerase chain (RT-PCR) for SARS-CoV-2 Ribonucleic acid (RNA), while those with severe COVID-19 additionally met at least one of the following conditions: (1) Shortness of breath or dyspnea, with a respiratory rate of 30 times/min, (2) Oxygen saturation (resting state) 93%, or (3) PaO2 / FiO2 300 mmHg.

Samples of patients with COVID-19 that were SARS-CoV-2 positive based on nucleic acid detection and showed the presence of lymphocyte subset population for approximately four weeks after onset, where lymphocyte subset analyses were performed at least twice within the four weeks duration, were included in this study. However, those samples that were SARS-CoV-2 nucleic acid positive and either lacked or showed the presence of only one lymphocyte subset population for approximately four weeks after onset were not considered for this study.

Patients with severe influenza were diagnosed based on Influenza: Diagnosis and Treatment (2019).[10] The diagnostic criteria were as follows: sustained high fever>3 days accompanied by severe cough, coughing up sputum or bloody sputum, or chest pain; rapid breathing rate, respiratory distress, and cyanosis of lips; change in consciousness such as unresponsiveness, drowsiness, restlessness, or convulsions; Severe vomiting, diarrhea, dehydration; presence of symptoms associated with pneumonia; exacerbation of pre-existing conditions and/or diseases; other clinical conditions requiring hospitalization. Patients with severe influenza A admitted to our hospital who met the above criteria were included in this study. Patients were excluded if an alternative diagnosis, such as the presence of influenza B, parainfluenza, respiratory syncytial virus, adenovirus, mycoplasma, chlamydia, legionella was determined.

Healthy controls who tested negative for human immunodeficiency virus (HIV) infection, hepatitis viral infections, systemic infections, connective tissue diseases, cancer, or any other abnormal tumor markers were considered.

Patients in four groups (non-severe COVID-19 and severe COVID-19, severe influenza A, and healthy controls) excluded minors younger than eighteen years old and pregnant women.

#### Data collection

The information related to age, gender, onset of symptoms, medical history of pre-existing comorbidities (hypertension, diabetes, chronic obstructive pulmonary disease (COPD), cardiovascular disease, chronic kidney disease, and/or autoimmune disease), severity assessment on admission, laboratory findings, and treatment regimen for each patient was extracted from electronic medical records.

#### Laboratory confirmation

Laboratory confirmation tests for COVID-19 were performed by Beijing Center for Disease Control and Prevention (CDC) and Beijing Ditan Hospital. Throat swab and/or sputum specimens were collected from all patients at admission and were tested the same day. Patients were confirmed to be positive for SARS-CoV-2 nucleic acid by real-time fluorescent reverse transcriptase-polymerase chain reaction assay, according to the WHO interim guidance for COVID-19.[11]

The diagnosis of severe influenza virus infection was confirmed by RT-PCR of nasopharyngeal swabs.[10]

#### Flow cytometric analyses

To explore the cellular immune response of patients with severe and non-severe COVID-19 and severe influenza A, peripheral blood samples (2mL each) were collected from healthy controls and all patients at week 1 (0–7 days), week 2 (8–14 days), week 3 (15–21 days), and week 4 (22–28 days) from the initial onset of symptoms, for analyses of complete blood cell counts and cell counts of lymphocyte subsets (CD4^+^ T cell, CD8^+^ T cell, B cell, and natural killer (NK) cell counts [cells/ and CD4^+^ T/CD8^+^ T ratio). Lymphocyte subsets were detected and counted by BD FACSCalibur flow cytometer (BD Biosciences, San Jose, CA, USA), and the subsets were characterized according to phenotypes of the corresponding CD antigens.

For T cell subset identification and counting, a Four ColorHuman CD3/CD8/CD45/CD4 Flow Kit was employed, according to manufacturer’s instructions. Briefly, 50 μL of the isolated peripheral blood mononuclear cells was incubated directly with the Four Color reagent consisting of a combination of RPE-Cy5.5 conjugated CD3 (Mouse IgG1), APC conjugated CD8 (Mouse IgG1), RPE conjugated CD45 (Mouse IgG2a), and fluorescein isothiocyanate (FITC)-conjugated CD4 (mouse IgG1) monoclonal antibodies for 15 min at 4°C in absolute counting tubes. Subsequently, 450 μL of red blood cell lysing solution (1xBD lysing solution) was added to the tubes before performing the flow cytometric analysis. The samples were then incubated at room temperature for 5 min prior to detection by BD FACS Calibur flow cytometer. Data were obtained and analyzed automatically by the MultiSET software.

### Statistical analyses

All analyses were performed using SPSS statistical software (version 26.0, IBM, Armonk, NY, USA). Categorical data are expressed in frequency or percentage, and statistical significance was determined by χ2 or Fisher’s exact test. Nonparametric variables are expressed as median with interquartile range (IQR). For continuous data, Mann-Whitney U test or Kruskal Wallis test was performed to compare variables between different groups and determine the statistical significance. In all analyses, *P* < 0.05 was statistically significant. The dynamic changes of different subsets of lymphocytes at different time periods are represented by boxplots.

## Results

### Demographic and clinical characteristics of patients with COVID-19 and severe influenza A

In our study, we retrospectively reviewed the clinical data of 110 healthy blood donors (healthy controls), 99 patients with COVID-19 and 43 patients with severe influenza A. The group with healthy controls (HC) comprised of 53 males (48%) and 57 females (52%). Among the 99 patients with COVID-19, 56 were men (56.6%) and 43 were women (43.4%). All the COVID-19 patients were divided into two groups, according to the abovementioned diagnostic criteria, and there were 80 non-severe cases (80.8%) and 19 severe cases (19.2%; Table 1). The median age of patients with non-severe and severe COVID-19 was 37 (30–49) years and 67 (53–74) years, respectively. The severe patient group had a higher age than the non-severe COVID-19 group. The median durations of hospitalization of patients with non-severe and severe COVID-19 were 4 (2, 7) and 5 (4.5, 10) days, respectively. All the patients with severe COVID-19 (n = 19, 100%) presented with fever, cough, and dyspnea. On the contrary, only a few patients with non-severe COVID-19 had fever (n = 58, 73%) and cough (n = 38, 48%). Notably, none of them had dyspnea. A total of 31 (31.3%) patients in both the COVID-19 groups had pre-existing chronic medical conditions, including diabetes (8 [11%]), hypertension (24 [32.9%]), COPD (2 [2.8%]), cardiovascular disease (2 [2.8%]), renal disease (1 [1.4%]), and autoimmune disorders (5 [6.9%]). Specifically, 19 patients (24%) with non-severe COVID-19 and 12 patients (63%) with severe COVID-19 presented with one or more of the pre-existing chronic medical conditions (Table 1). One patient in the severe COVID-19 group died on day 28 after the onset of the disease.

**Table 1.**
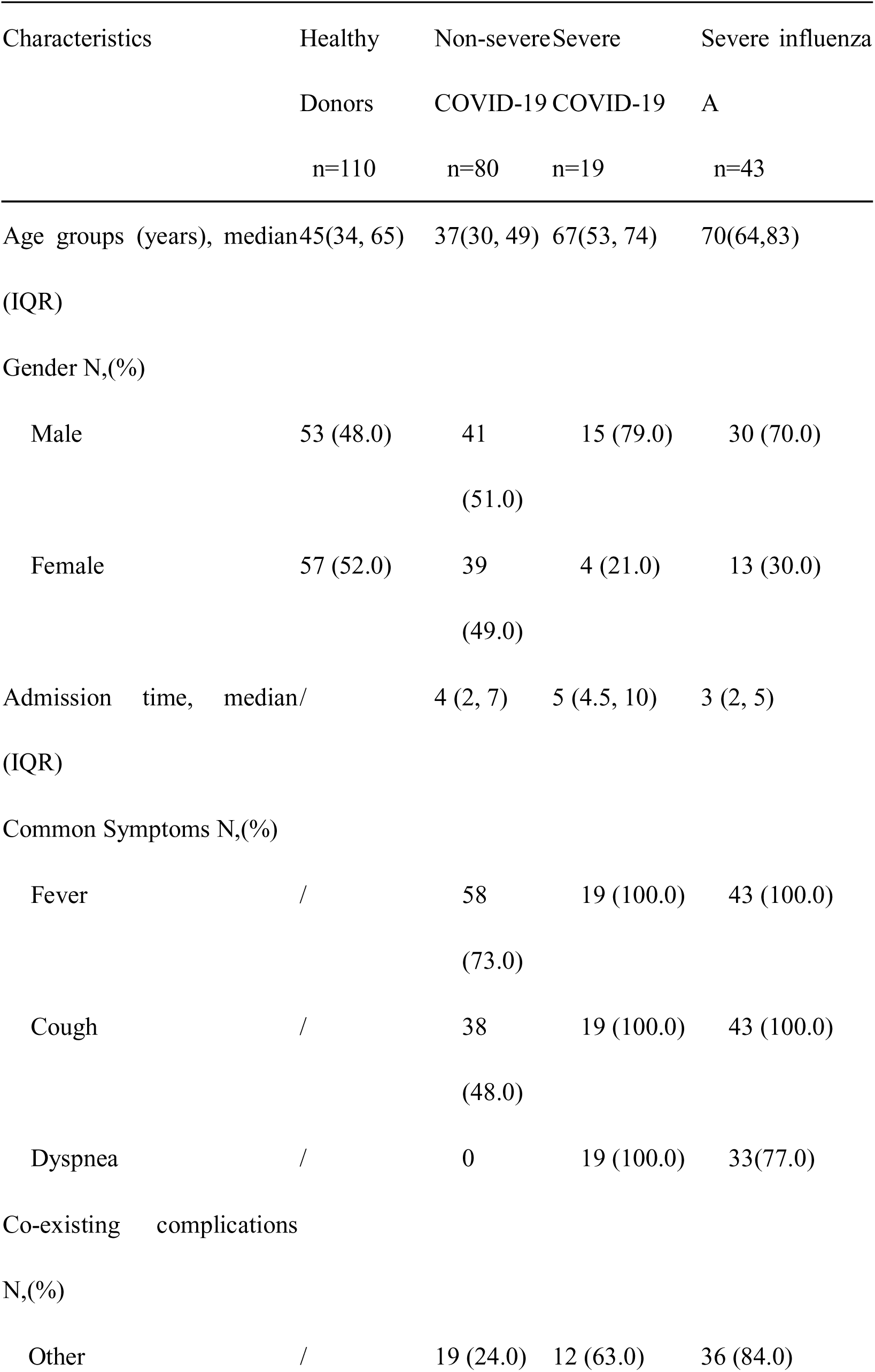
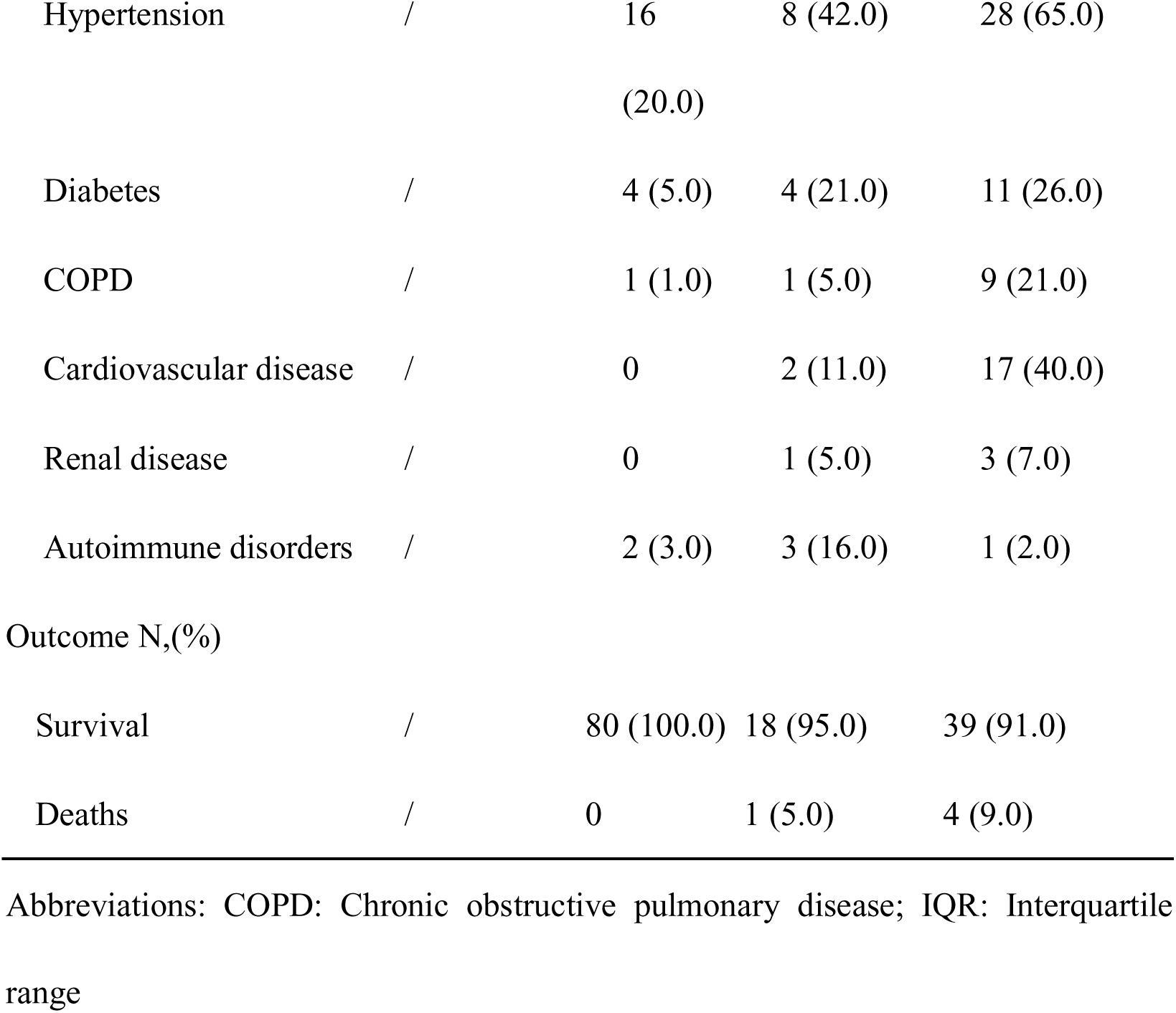
Demographic and clinical characteristics of different study groups Characteristics Healthy

Similar to the group with severe COVID-19, the median age of patients with severe influenza A infection was higher (70 years) than that of those of healthy controls (45 years). Notably, the difference in age was found to be statistically significant (*P* < 0.05) between the groups. All patients with severe influenza A infection (n = 43, 100%) presented with fever and cough. However, only 33 patients (77%) had dyspnea. Out of 43 patients with severe influenza A, 4 (9%) died and 39 (91%) survived (Table 1).

### Variations in leukocyte populations in COVID-19 and severe influenza A

We analyzed different variations in leukocyte subpopulations of patients with non-severe and severe COVID-19 and severe influenza A during the first week of illness. The absolute counts of total white blood cells (WBCs) of patients with non-severe (5.34 ×10^9^/L, *P* < 0.001) and severe COVID-19 (3.92 ×10^9^/L, *P* < 0.0001) were significantly lower than those of the healthy donors (6.01 ×10^9^/L; Table 2). Moreover, a statistically significant difference was observed between WBC counts of patients with non-severe and that of those with severe COVID-19 (*P =* 0.049). Further, there was a significant difference between the WBC counts of patients with severe influenza A and those of patients with severe COVID-19 (*P =* 0.027).

**Table 2.**
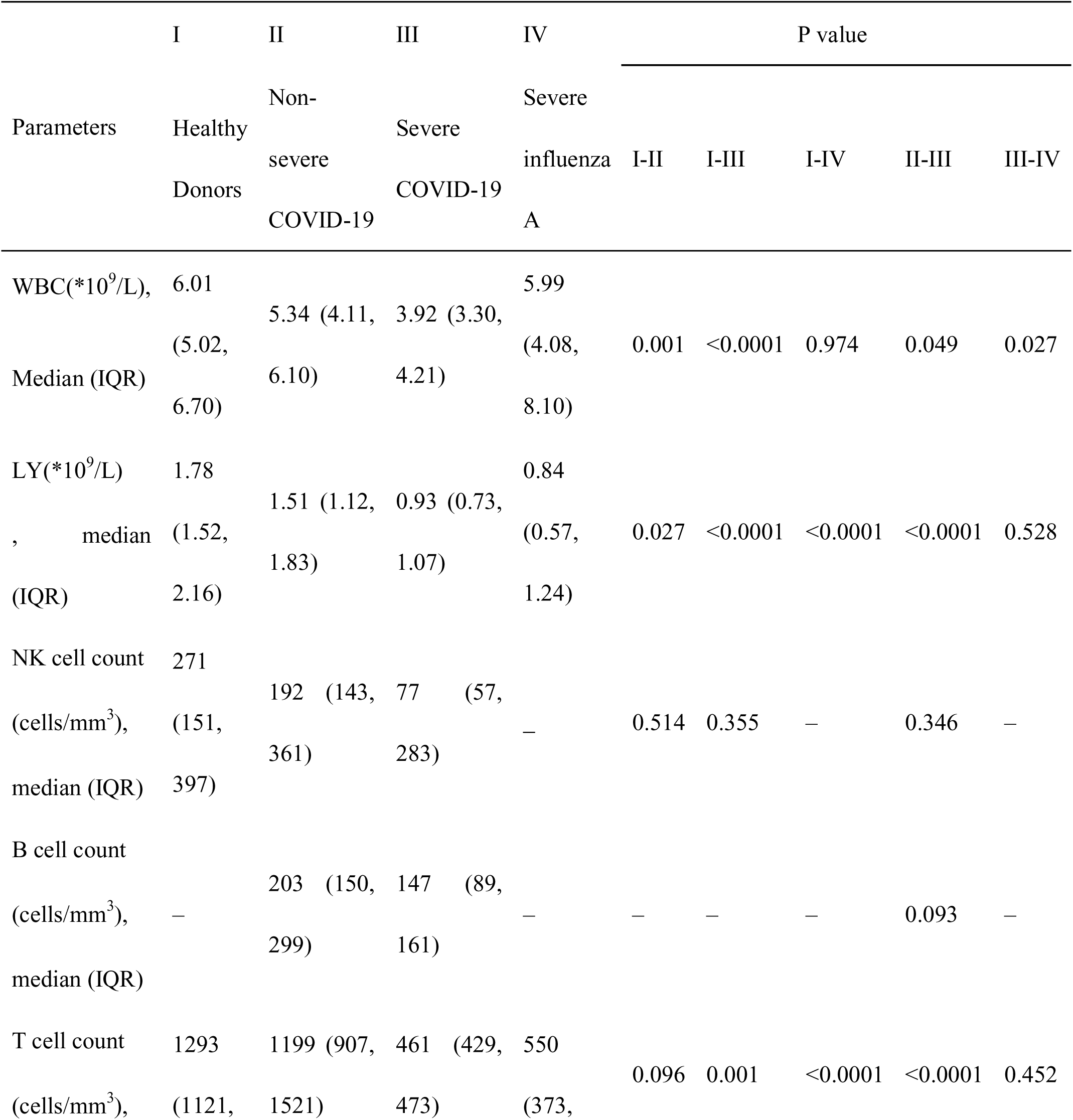
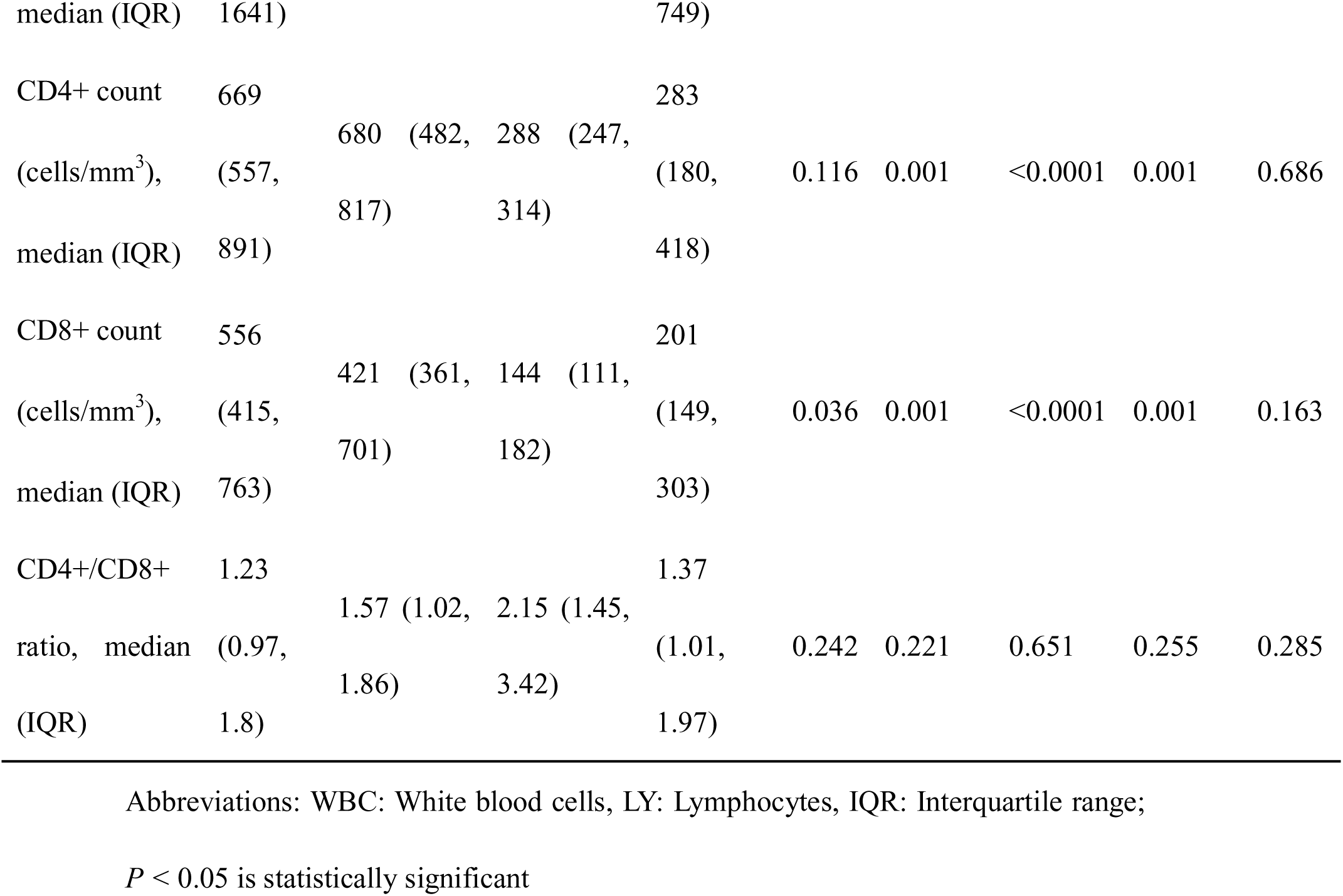
Summary of comparison of cell counts of total WBCs, total lymphocytes, and lymphocyte subsets in patients with non-severe COVID-19, severe COVID-19, and severe influenza A with the first week of illness.

Expectedly, the absolute counts of lymphocytes in healthy donors (1.78 ×10^9^/L) were significantly higher than those of patients with non-severe COVID-19 (1.51 ×10^9^/L, *P =* 0.027), severe COVID-19 (0.93 ×10^9^/L, *P* < 0.0001), and severe influenza A (0.84 ×10^9^/L, *P* < 0.0001) during the first week of illness (Table 2). Further, there was a significant difference in the absolute lymphocyte counts of patients with non-severe and severe COVID-19 (*P* < 0.0001).

### Distinct alterations in peripheral lymphocyte subsets in COVID-19 and severe influenza A

To further determine variations in different lymphocyte subsets in peripheral blood samples of patients with non-severe COVID-19, severe COVID-19, and severe influenza A during the first week of illness, we performed flow cytometric analysis to enumerate total T cell population, CD4^+^ and CD8^+^ T cell subsets, B cells and NK cells. Similar to the findings of the absolute counts of total lymphocytes, patients with severe COVID-19 and severe influenza A had a significantly lower number of total T cells (*P =* 0.001 and *P <*0.0001, respectively), CD4^+^ T cells (*P =* 0.001 and *P <*0.0001, respectively), and CD8^+^ T cells (*P =* 0.001 and *P <*0.0001, respectively) than healthy controls.

Notably, there was a significant difference between the CD8^+^ T cell subset (*P =* 0.036) counts of patients with non-severe COVID-19 and those of healthy donors. The differences in the total T cell population (*P <*0.0001), CD4^+^ T cell subset (*P =* 0.001), and CD8^+^ T cell subset (*P =* 0.001) of patients with non-severe and severe COVID-19 were statistically significant (Table 2).

In addition, the median of CD4^+^/CD8^+^ ratio in non-severe COVID-19, severe COVID-19 and severe influenza group showed no different compared to HCs (*P* = 0. 242, *P* = 0.221 and *P* = 0.651, respectively). And at the same time, the median NK cells of non-severe and severe COVID-19 patients showed no difference compared to HCs (*P* = 0. 514 and *P* = 0.355, respectively). There was no significant difference in B cells between the severe and non-severe COVID-19 groups (*P* = 0.093). (Table 2)

### Dynamic changes in leukocyte and lymphocyte subset counts with disease progression and assessment of severity

As shown in Table 2, total WBCs, the absolute lymphocytes, total T cells, CD4^+^ and CD8^+^ T cell subsets in the peripheral blood samples of all patients were found to be lower than those of healthy donors at the time of disease onset. To further determine significant changes in the immune cell subsets of patients with non-severe COVID-19, severe COVID-19, and severe influenza A, with disease evolution, we compared the absolute counts of leukocytes, total lymphocytes, and lymphocyte subsets of the three patient groups at weeks 1, 2, 3, and 4.

Unlike other groups, in patients with severe COVID-19, leukocyte counts gradually increased from week 1 to week 4, despite being lower than the normal. Notably, the variations in leukocyte counts were found to be statistically significant. (W1-W2: *P =* 0.013, W1-W3: *P =* 0.003, W1-W4: *P =* 0.001). A similar trend was observed with total lymphocyte count, which sharply increased at week 3, showing a significant difference compared to those at weeks 1 and 2 (W1–W3: *P =* 0.013, W1-W4: *P =* 0.002, W2-W3: *P =* 0.001) (**Fig. 1 A, B**). On the other hand, a significant and a drastic increase was noted in the cell counts of total T cell and CD4^+^ T cell subset at week 4 when compared with those of weeks 1 and 2. (W1-W4: *P =* 0.034, W2-W4: *P =* 0.003; W1-W4: *P =* 0.020, W2-W4: *P =* 0.001)(**Fig. 1 C- E**).

**Fig. 1.**
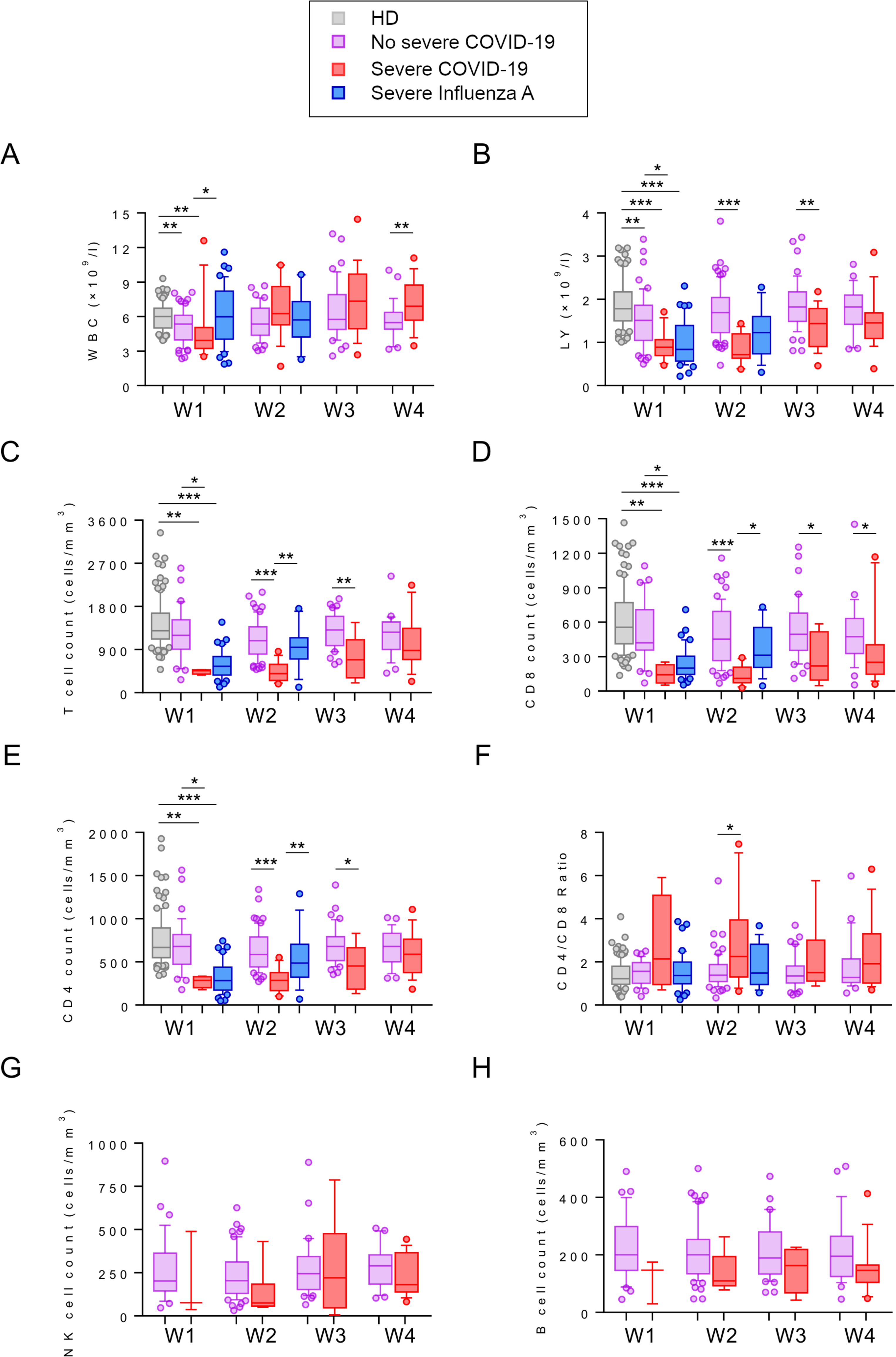
Dynamic variations in cell counts of total WBCs, total lymphocytes, and lymphocyte subsets in patients with non-severe COVID-19, severe COVID-19, and severe influenza A at weeks (W) 1, 2, 3, and 4 during the course of illness. **(A–B)** Box plots illustrating percentages of total WBCs and total lymphocytes in different groups at different times. Kruskal–Wallis test followed by Dunn’s multiple comparisons test or one-way analysis of variance (ANOVA) test followed by Tukey’s multiple comparisons test was performed to determine significant differences between the groups. **(C–H)** Box plots illustrating total cell counts of lymphocyte subsets in different groups at different times. Spearman’s nonparametric test was used to test for correlations. ** P< 0.01, *** P< 0.001 were considered significant.

Consistent with the findings of patients with severe COVID-19, the leukocyte counts of patients with non-severe COVID-19 were significantly higher in week 3 than those of week 1 and week 2 (W1-W3: *P =* 0.005, W2-W3: *P =* 0.044). A similar tendency was seen with lymphocyte count, which showed a sharp significant increase at week 3 compared with that seen at week 1 (W1-W3: *P =* 0.004). Interestingly, a significant change in total T cell count was observed at week 3, suggesting a marked increase in total T cell population when compared with that observed at week 2 (W2-W3: *P =* 0.045)(**Fig. 1 A-C**). The differences in the absolute counts of total lymphocytes, total T cells, CD4^+^ and CD8^+^ T cell subsets were found to be significant between the severe and non-severe COVID-19 groups at all time points (weeks 1 through 4), although they were visibly lower than those of healthy controls (**Fig. 1 B-E**). When compared with healthy controls, no significant changes were seen in B cell populations and CD4^+^/CD8^+^ ratios of patients with severe and non-severe COVID-19 during the four weeks following the infection (**Fig. 1 F, H)**. Interestingly, the NK cell counts of patients with severe and non-severe COVID-19 gradually increased from weeks 2 to 4. These changes were found to be statistically significant (W2-W4: *P =* 0.011 and *P =* 0.042, respectively) (**Fig. 1 G)**. On the contrary, in patients with severe influenza A infection, significant changes were only evident in the cell counts of total T cells, CD4^+^ and CD8^+^ T cell subsets (W1-W2: *P =* 0.003, *P =* 0.006, and *P =* 0.030, respectively) (**Fig. 1 C-E**).

## Discussion

In this study, we aimed to analyze the role of lymphocyte subsets in the immunopathogenesis of COVID-19 and severe influenza A, and examined the clinical significance of their alterations, especially in determining the prognosis and recovery duration. Our analyses revealed significant dynamic variations in total lymphocytes and lymphocyte subsets, which get activated in the early stages of COVID-19 and severe influenza A infections, and further demonstrated that severe immune injury tended to be more prominent in patients with severe form of the disease. The recovery rate of patients with severe COVID-19 was comparatively longer than those who received immediate antiviral treatment for severe influenza A and those with non-severe COVID-19.

We retrospectively reviewed the clinical data of 99 patients who were confirmed to have COVID-19, 43 patients with severe influenza A and 110 healthy blood donors who were previously recruited in 2018. All the patients with COVID-19 were divided into two groups, according to the abovementioned diagnostic criteria, including 80 non-severe cases (80.8%) and 19 severe cases (19.2%). Several reports and studies have clearly indicated that older or elderly people are more prone to COVID-19 as their immune systems are likely to get overwhelmed by infections due to their advanced age. Similarly, the elderly who are 65 years or older are particularly at risk for influenza infection, hospitalization, and death due to influenza-related complications, such as pneumonia.[12] In our study, the median age of patients with non-severe COVID-19 was 37 years, while that of those with severe COVID-19 was 67 years. Consistent with the previous studies, our data indicate that the ages of the severe patient group are higher than those of the non-severe COVID-19 group. Further, our study showed that the median age of patients with severe influenza A infection was higher (70 years) than that of those with non-severe COVID-19 (37 years) and healthy controls (45 years). Notably, this difference in age was found to be statistically significant (*P* < 0.05) among the groups, which suggests that the elderly people represent a large at-risk population.

The WHO-China joint report on COVID-19 provided a comprehensive symptomatology of COVID-19 (n = 55,924)[13]. A previous study showed that patients with COVID-19 present with pyrexia in 85% of cases during their illness course, but only 45% are febrile on early presentation. In addition to cough (67.7%) and sputum (33.4%), respiratory symptoms, such as dyspnea, sore throat, and nasal congestion were reported to be present in 18.6%, 13.9%, and 4.8% of cases, respectively. Both COVID-19 and influenza present with common clinical manifestations including fever, cough, rhinitis, sore throat, headache, dyspnea, and myalgia. [7,14,15]In our study, all the patients with severe COVID-19 presented with fever, cough, and dyspnea, whereas only 73% and 48% of patients with non-severe COVID-19 had fever and cough, respectively. Similar to the severe COVID-19 group, all patients with severe influenza A infection presented with fever and cough. However, only 77% of the patients had dyspnea. These clinical manifestations of COVID-19 and influenza infections were consistent with other studies. As previously reported the mortality rate also increases in patients with additional comorbidities.[3] Specifically, 63% and 84% patients with severe COVID-19 and severe influenza A, respectively, presented with one or more pre-existing chronic medical conditions. While the mortality rate was 5% due to severe COVID-19, it was even higher (9%) for severe influenza A.

Lymphocytes and their subsets play a crucial role in maintaining immune homeostasis and inflammatory response in the host. As in case of immune disorders and other infections, a viral infection impairs the host’s immune defenses and results in decreased levels of lymphocytes and their subsets.[16,17] Lymphocyte subsets, namely CD4^+^ T cells, CD8^+^ T cells, B cells, and NK cells are primarily involved in the humoral and cytotoxic immunity against viral infection. Therefore, this necessitates the need to understand the mechanism of reduced blood lymphocyte levels and characterize the dynamic alterations of lymphocyte subsets to provide novel insights for an effective treatment and prognosis of COVID-19. As lymphopenia is frequently observed during the initial stages of respiratory viral infection.[3,18] We identified and analyzed different variations in leukocytes, lymphocytes, and lymphocyte subsets in patients with non-severe and severe COVID-19 and severe influenza A during the first week of illness. Our results showed that the absolute counts of total WBCs and lymphocytes of patients with non-severe (5.34 ×10^9^/L, *P* < 0.001 and 1.51 ×10^9^/L, *P =* 0.027, respectively) and severe COVID-19 (3.92 ×10^9^/L, *P* < 0.0001 and 0.93 ×10^9^/L, *P* < 0.0001, respectively) were significantly lower than those of healthy donors (6.01 ×10^9^/L and 1.78 ×10^9^/L, respectively). Further, the total lymphocyte counts of patients with severe influenza A were comparatively lower than those of healthy donors, (0.84 ×10^9^/L, *P* < 0.0001). These findings are corroborated by a previous study which hypothesized that the virus might directly infect lymphocytes resulting in their apoptosis, thus leading to causing a sharp decline in total lymphocyte population and subsequent lymphopenia. Moreover, lymphocytes express the coronavirus receptor angiotensin-convertingenzyme 2(ACE-2), and therefore are a direct target for the virus.[19] Another retrospective study suggested that lymphopenia might be one of the predictive factors for progression to respiratory failure during early stages following Middle East Respiratory Syndrome coronavirus MERS-Cov) infection.[20] Giving further strength to our study, a study by Geng et al. demonstrated the decline in the populations of T lymphocytes and their subsets, after influenza A virus infection, to be positively correlated with prognosis.[21] We further performed flow cytometric analysis to enumerate total T cell population, CD4^+^ and CD8^+^ T cell subsets, B cells and NK cells in patients with non-severe COVID-19, severe COVID-19, and severe influenza A to determine significant changes in different lymphocyte subsets during their first week of illness. Our study demonstrated that patients with severe COVID-19 and severe influenza A had a significantly lower number of total T cells (*P =* 0.001 and *P <*0.0001, respectively), CD4^+^ T cell subsets (*P =* 0.001 and *P <*0.0001, respectively), and CD8^+^ T cell subsets (*P =* 0.001 and *P <*0.0001, respectively) than healthy controls. Further, significant differences in total WBCs (*P =* 0.049), total lymphocytes (*P* < 0.0001) total T cell population (*P* < 0.0001), CD4^+^ T cell subsets (*P =* 0.001), and CD8+ T cell subsets (*P =* 0.001) were observed between patients with non-severe and severe COVID-19. Notably, there was a significant reduction in CD8^+^ T cell subsets (*P =* 0.036) in patients with non-severe COVID-19 compared with healthy controls. This indicates a more obvious change in CD8^+^ T cell subsets than in other lymphocyte subsets following SARS-CoV-2 infection. Therefore, our results reiterate the fact that CD8+ T cell responses play a major role in antiviral immunity.[22] Taken together, lymphopenia was common in the patients with COVID-19, indicating a significant impairment in the host’s immune system following SARS-CoV-2 and influenza A infections. Our findings are in line with other studies which also detected these alterations in patients with pneumonia caused by MERS-CoV and Severe acute respiratory syndrome coronavirus (SARS-CoV).[20, 23] In addition, a significant reduction in both CD4^+^ T cells and CD8+ T cells were specifically observed in patients with severe COVID-19 and severe influenza A. Therefore, this indicates a more severe immune insult in patients with the severe form of the disease. Consequently, the alteration would be more profound, and leads to adverse clinical outcome in these patients. Thus, lymphocytes and their subsets, especially CD8^+^ T cells, might be a potential predictor for disease severity and clinical efficacy in COVID-19.

Next, we focused on the dynamics of T lymphocytes and their subsets, which played a vital role in cellular immune responses. We compared the absolute counts of leukocytes, total lymphocytes, and lymphocyte subsets of the non-severe and severe COVID-19 patient groups at weeks 2, 3, and 4 with that of those observed during the initial stages of infection. The total leukocyte, lymphocyte, and T cell counts significantly improved at week 3 in patients with non-severe COVID-19. Lymphocyte also recovered markedly at week 3 in severe COVID-19 patients. However, T cell and CD4^+^ T cell subset population significantly increased at week 4 in patients with severe COVID-19. Our results are consistent with a previous study by He et al. that showed a sharp decline (below normal) in the cell counts of CD45^+^, CD3^+^ T cell subsets, CD4^+^ T cell subsets, and CD8^+^ T cell subsets during the first week of SARS-Cov infection; their values further declined during the second week before increasing during the third week and returning to normal by the fifth week. Moreover, CD4^+^ T and CD8^+^ T cell counts were found to be extremely low in critically ill and deceased patients.[23] Taken together, the alterations in lymphocytes and their subsets gradually improved at later time points in patients with COVID-19. Collectively, our results indicate that the recovery duration of patients with severe COVID-19 is longer than those with the mild form of the disease.

Our study results further revealed a noticeable difference in the time taken for the cell counts to improve among the severe and non-severe COVID-19 and the severe influenza groups. The cell counts of total lymphocytes and their subsets recovered only around week 4 in severe COVID-19; the recovery time was almost delayed by a week compared with those having non-severe COVID-19. On the contrary, the cell counts of total lymphocytes and their subsets in patients with severe influenza A increased and improved drastically at week2; this rapid recovery rate could be attributed to the early initiation of treatment with the neuraminidase inhibitor, oseltamivir, or peramivir, which interfere with virus release from host cells by blocking the viral nucleic acid function, thus preventing infection of new host cells.[24]

NK cells are cytotoxic innate lymphocytes that play an important role in controlling the viral burden. NK cell responses can be specific, and they interact with both innate and adaptive immune cells to coordinate appropriate antiviral responses.[25] Although we observed a reduction in NK cell counts during the initial stages of infection in patients with severe and non-severe COVID-19, their recovery rate (cell counts improved at week 3) showed a similar trend as that of the lymphocytes in both the groups. One plausible reason for this observation could be that the NK cell cross-talk might have got suppressed following the virus attack, which might have further led to impairment of CD4^+^ T cell responses and their effects on CD8^+^ T cells. This is further supported by a finding from this study which demonstrated a significant reduction in CD4^+^ T cell subsets, especially in patients with severe COVID-19. Given the role of B cells in adaptive immune response, we did not observe any significant changes in total B cell population in all patients with COVID-19 during the course of illness. This could possibly be attributed to the poor activation of the adaptive immune response against the virus. Moreover, the lack of detectable virus during stage I (asymptomatic incubation period)of the SARS-CoV-2 infection might have failed to elicit protective B cell immunity.[26] Therefore, future research focusing on strategies to enhance the specific adaptive immune responses during the incubation and non-severe stages of the SARS-CoV-2 infection could significantly boost the B cell population, preventing the progression to the stage III severe respiratory symptomatic stage with high viral load. Furthermore, studies identifying specific immune components or functions that limit protective immunity to virus infection are warranted.

Our study has several limitations. This study was retrospective, small single-center study and only included a small sample of 99 patients with COVID-19 admitted to Beijing Ditan Hospital, which may confound the results and potentially introduce selection bias. This may limit the generalizability of the study. In addition, inconsistencies in time periods between illness onset and admission might have led to missing data which could result in observation biases in the dynamic variations in immune cells.

## Conclusions

Collectively, our study indicates that lymphopenia might be associated with disease severity and suggests the plausible role of lymphocyte subsets in disease progression, which in turn affects prognosis and recovery duration in patients with severe COVID-19 and influenza A. The dynamic alterations in lymphocytes and their subsets provides valuable insights into the immunopathogenesis of disease progression in COVID-19.

## Data Availability

The datasets used and/or analysed during the current study are available from the corresponding author on reasonable request.

## Abbreviations

COVID-19: Coronavirus disease-19
WHO: World Health Organization
SARS-CoV-2: Severe acute respiratory syndrome coronavirus 2
CT: Computed tomography
RT-PCR: Reverse transcription-polymerase chain reaction
RNA: Ribonucleic acid
HIV: human immunodeficiency virus
COPD: chronic obstructive pulmonary disease
CDC: Center for Disease Control and Prevention
NK: natural killer
IQR: interquartile range
HC: healthy controls
WBC: White blood cells
LY: Lymphocytes
ACE-2: angiotensin-convertingenzyme 2
MERS- CoV: Middle East Respiratory Syndrome coronavirus
SARS-CoV: Severe acute respiratory syndrome coronavirus

## Declarations

### Ethics approval and consent to participate

This study was approved by the Ethics Committee of Beijing Ditan Hospital (No. 202000601). The need for individual consent was waived because of the retrospective nature of the study.

### Consent for publication

Not applicable.

### Availability of data and material

The datasets used and/or analysed during the current study are available from the corr esponding author on reasonable request.

### Authors’ contributions

FZ and ZC were responsible for the study concept and design. They had complete access to all the data in this study and take responsibility for data integrity and accuracy of the data analysis. FQ and YS contributed to the writing of this manuscript. FQ, GG, YX, AW, SW, XM, TZ and XG contributed to the sample acquisition of patients, diagnosis, and treatment. FQ and YS contributed to data collection, data interpretation, and analysis of the results. YH and MC conducted the statistical analyses. All authors reviewed and approved the final manuscript.

## Acknowledgments

We acknowledge all health-care workers involved in the diagnosis and treatment of patients in Beijing Ditan Hospital, Capital Medical University.

